# A multicenter study to assess the performance of the point-of-care RT-PCR Cobas SARS-CoV-2 & Influenza A/B nucleic acid test for use on the Cobas Liat system in comparison with centralized assays across healthcare facilities in the United States

**DOI:** 10.1101/2024.09.06.24313202

**Authors:** Elissa M. Robbins, Rasa Bertuzis, Ho-Chen Chiu, Lupe Miller, Christopher Noutsios

**Author notes:** **Corresponding author:** Elissa M. Robbins.

## Abstract

Respiratory diseases can share many of the same symptoms, highlighting the need for timely and accurate differentiation to facilitate effective clinical management and reduce transmission. Compared with centralized testing, molecular point-of-care tests (POCT) can provide a faster time to result.

We evaluated the RT-PCR POCT Cobas^®^ SARS-CoV-2 & Influenza A/B qualitative assay for use on the Cobas Liat^®^ system (the POC SARS-CoV-2 & Influenza A/B test) in nasal and nasopharyngeal swab samples from 10 diverse healthcare facilities in the US. A composite comparator design consisting of three centralized tests was used to analyze SARS-CoV-2, whilst performance versus a single centralized test was used for analysis of influenza A and B. Evaluations included performance stratified by sample type (prospective/retrospective and nasal/nasopharyngeal [paired by subject]), collection method (self/healthcare worker-collected [alternated and approximately balanced], symptom status (symptomatic/asymptomatic), and SARS-CoV-2 vaccination status, as well as assay inclusivity and system ease of use.

A total of 2,247 samples were tested. For SARS-CoV-2, the overall percent agreement (OPA) was 98.8% (95% CI: 97.9, 99.3) in nasal swab samples and 99.0% (95% CI: 98.2, 99.4) in nasopharyngeal swab samples. Regression analysis showed that Ct values from paired nasal and nasopharyngeal swab samples were highly correlated (correlation coefficient 0.83). The OPA was ≥99.5% (sample type dependent) and 100.0% for influenza A and B, respectively. The POC SARS-CoV-2 & Influenza A/B test was easy to use.

These results support the use of the POCT in various sample types and by various operators in the intended-use setting.

**Summary statement/importance:** This study highlights the benefits of RT-PCR POCT, namely comparable performance to centralized testing in multiple sample types and ease of use. Utilizing assays such as the POC SARS-CoV-2 & Influenza A/B test may improve timely differentiation of respiratory diseases that share similar symptoms.

## Introduction

More than 4 years since the start of the coronavirus disease 2019 (COVID-19) pandemic, much of the world has returned to normality, yet COVID-19 continues to cause severe illness and mortality in the United States (US) and elsewhere(1, 2). This applies particularly for the immunocompromised, who are at risk of severe outcomes (3) and comprise approximately 3% of the US adult population(4). Disease activity is year-round, but seasonal spikes occur from November to April(5), coinciding with circulation of other respiratory viruses, such as influenza(6). Respiratory diseases such as COVID-19 and influenza share similar symptoms(7), highlighting the need for timely and accurate differentiation of the causative agent to help facilitate effective clinical management and reduce onward transmission.

The current choice of testing for severe acute respiratory syndrome coronavirus 2 (SARS-CoV-2) is setting dependent(8), but standard laboratory-based (i.e., centralized) nucleic acid amplification tests are the mainstay of diagnosis(9). Numerous antigen or molecular point-of-care tests (POCT) are commercially available(10, 11), offering the benefit of a faster time to result(12). Both POCT formats have lower performance compared with centralized testing, particularly sensitivity (13) — a secondary analysis of antigen POCT found the performance to be commonly lower than stated within manufacturers’ instructions for use (14) — but molecular POCT have relatively higher performance in comparison with antigen POCT, with pooled sensitivity values of 93% (99% specificity) versus 71% (99% specificity), respectively(13). Molecular POCT can provide actionable results in as little as 20 minutes depending on the assay and workflow(11).

Influenza is a significant cause of severe illness and death(15–17), resulting in a substantial clinical burden to patients and the healthcare system alike(18). Molecular POCT for the detection of influenza A/B subtypes, and other respiratory viruses such as respiratory syncytial virus, have been available for several years(11, 12, 19–22). Clinical practice guidelines from the Infectious Diseases Society of America recommend that antivirals are initiated for certain patient groups as soon as possible regardless of illness duration, or within 48 hours of illness onset(23), supporting the use of rapid molecular POCT for influenza testing in preference to centralized laboratory testing. In the hospital setting, an interventional study of a molecular POCT for influenza found significant improvements in isolation practices and reductions in length of stay compared with centralized testing(24), whilst a modeling study found that the introduction of POCT could reduce time to diagnosis, hospital stay, and in-hospital costs(25). Molecular POCT for influenza can generate sensitivity estimates of ≥96–100% in real-world settings(22, 26).

The primary objective of this study was to evaluate the real-world clinical performance of the POC Cobas SARS-CoV-2 & Influenza A/B qualitative assay for use on the Cobas Liat system (herein referred to as the POC SARS-CoV-2 & Influenza A/B), a multiplexed RT-PCR test combining measurement of SARS-CoV-2 and influenza subtypes, in reference to comparator centralized PCR tests for SARS-CoV-2 and influenza A and B viruses. Sample types were nasal and nasopharyngeal swab specimens from individuals suspected of respiratory viral infection consistent with COVID-19, and asymptomatic individuals exposed to COVID-19, who presented at healthcare facilities across the US during the COVID-19 public health emergency. Secondary objectives were to evaluate the influence of collection method and the ease of use of the Cobas Liat system.

## Materials and methods

### Sites

Testing was conducted at 11 sites: 10 geographically diverse healthcare facilities that were representative of intended-use sites (i.e., POC settings, such as emergency departments, urgent care, pediatric and primary care clinics, and drive-through COVID-19 testing sites), and one reference laboratory for centralized testing. After testing using the POC SARS-CoV-2 & Influenza A/B test at the healthcare facilities, samples were shipped to the reference laboratory for analysis with the comparator assays (see below).

Healthcare facilities were selected based on sample availability, ability to provide adequate resources and operators, ability to adhere to good clinical practice, and with the aim of ensuring representation of the demographic diversity of the US population. Not all healthcare facilities performing testing were certified under Clinical Laboratory Improvement Amendments (CLIA) regulations to perform waived testing, but all facilities met the requirements for CLIA-waived settings (i.e., intended-use settings using untrained non-laboratory personnel). All healthcare facilities performed both sample collection and POC testing. POC operators at the 10 healthcare facilities (n=30 operators in total) had limited or no laboratory training and were representative of typical test operators in CLIA-waived settings (e.g., nurses, nursing assistants, and medical assistants). Operators at the reference laboratory were blinded to the results generated at the healthcare facilities.

### Study population

The symptomatic group comprised samples from individuals with suspected respiratory viral infection consistent with COVID-19 (see Signs/symptoms, Supplementary material). The asymptomatic group comprised individuals with self-reported ‘recent’ exposure (timeframe undefined) to SARS-CoV-2-positive individuals or any other reasons to suspect COVID-19. Patient clinical information was gathered, including, but not limited to, antiviral usage for up to 7 days prior to and on the day of sample collection, gender, race/ethnic group, and age. Individuals presenting at the healthcare facilities were considered for study inclusion, subject to the eligibility criteria (see Eligibility criteria, Supplementary material).

### Study design

The clinical performance of the POC SARS-CoV-2 & Influenza A/B test (27) was evaluated by comparing results to a composite comparator method for the SARS-CoV-2 analyte using the following three highly sensitive centralized testing assays: Cobas SARS-CoV-2 qualitative assay for use on the Cobas 6800/8800 systems (herein referred to as the 6800/8800 SARS-CoV-2 test)(28); Cobas SARS-CoV-2 & Influenza A/B qualitative assay for use on the Cobas 6800/8800 systems (herein referred to as the 6800/8800 SARS-CoV-2 & Influenza A/B test)(29); and Hologic^®^ Aptima^®^ SARS-CoV-2 Assay(30).

The clinical performance for the influenza A/B analyte components of the POC SARS-CoV-2 & Influenza A/B test was evaluated against a single centralized testing assay, the 6800/8800 SARS-CoV-2 & Influenza A/B (29).

Samples that generated invalid results were repeated where sample volume permitted additional testing; samples generating a second invalid result upon retesting were reported as invalid. Interrogation of discrepant samples was not performed, though selected samples were further retested with the POC SARS-CoV-2 & Influenza A/B test for exploratory purposes.

### Sample types and collection

Sample types included fresh and frozen nasal and nasopharyngeal swab samples, collected in the same media formulations but under different brand names: Copan Universal Transport Medium (UTM) or BD Universal Viral Transport (UVT) medium.

#### Prospective fresh samples (UTM)

One nasal swab and one nasopharyngeal swab sample were collected from each study participant. A nasal swab of both nostrils was first collected either by the healthcare worker (HCW), herein referred to as ‘HCW-collected’, or by the study participant under instruction from the HCW, herein referred to as ‘self-collected’, as per manufacturer’s instructions.(31) A nasopharyngeal swab was then collected by HCWs from the same study participant to generate paired samples. If a nasopharyngeal swab had already been collected (using one nostril) as part of the healthcare facility’s standard of care, the study nasopharyngeal swab was taken from the opposite nostril.

All prospective samples were collected from patients presenting to healthcare facilities during February to June 2022, characteristic of the 2021/2022 winter respiratory season.

#### Retrospective frozen samples (archived or purchased from external vendors; UTM or UVT)

The low influenza prevalence in the prospective population was anticipated because of reduced international travel, new respiratory care paradigms, increased public health awareness, and use of interventions (such as masking policies or social distancing) that occurred during the COVID-19 pandemic and study enrollment period, in which influenza A virus had been the dominant virus type in circulation.(32) Therefore, frozen samples positive for influenza A virus and for influenza B virus were used to supplement the prospectively collected fresh samples. These retrospective samples were collected in the US during the 2013–2014, 2014–2015, and 2019–2020 influenza seasons. Baseline demographic data and patient characteristics related to these samples were unavailable.

Retrospective samples with known SARS-CoV-2 status, collected between March 29, 2021 and May 26, 2021, were also included in the SARS-CoV-2 analyses.

Whilst multiple freeze-thaw cycles have a limited effect on the viral titer(33), there is the potential for samples that were initially low-titer positives to appear negative following storage. Internal Roche data support the stability of viral titer for up to three freeze-thaw cycles, so retrospective samples were only eligible for inclusion if they had undergone no more than two freeze-thaw cycles. Retrospective positive and negative samples were blended in a standard POCT workflow and assessed alongside prospectively collected samples.

### Details of assays and instruments

#### POC SARS-CoV-2 & Influenza A/B test (POC RT-PCR)

The Cobas Liat analyzer is for *in vitro* diagnostic use(27). The analyzer automates all nucleic acid amplification test processes, including target enrichment, inhibitor removal, nucleic acid extraction, reverse transcription, DNA amplification, real-time detection, and result interpretation in approximately 20 minutes(27). The Cobas Liat system comprises the Cobas Liat analyzer in conjunction with the Cobas Liat assay tubes(27).

The POC SARS-CoV-2 & Influenza A/B test utilizes a single-use disposable assay tube that contains all the reagents necessary for the detection of SARS-CoV-2 and hosts the sample preparation and PCR processes(34). This multiplex real-time RT-PCR test is intended for the simultaneous rapid *in vitro* qualitative detection and differentiation of SARS-CoV-2, influenza A, and influenza B virus RNA in healthcare provider-collected nasal and nasopharyngeal swab samples and self-collected nasal swab samples (collected in a healthcare setting with instruction by a healthcare provider) from individuals suspected of respiratory viral infection consistent with COVID-19(34).

#### 6800/8800 SARS-CoV-2 & Influenza A/B test (centralized testing method [CTM] 1)

The Cobas 6800/8800 systems consist of the sample supply module, the transfer module, the processing module, and the analytic module(29). Automated data management is performed by the Cobas 6800/8800 software, which assigns results for all tests. Results are available within less than 3.5 hours after loading the sample on the system.(29) The positive SARS-CoV-2 result is defined as positive on the SARS-CoV-2 channel (Target 2) and/or the pan-sarbeco channel (Target 3).

The 6800/8800 SARS-CoV-2 & Influenza A/B test is an automated multiplex real-time RT-PCR assay intended for simultaneous qualitative detection and differentiation of SARS-CoV-2, influenza A virus, and/or influenza B virus RNA in healthcare provider-collected nasal and nasopharyngeal swab samples and self-collected nasal swab samples (collected in a healthcare setting with instruction by a healthcare provider) from individuals suspected of respiratory viral infection consistent with COVID-19(29).

#### 6800/8800 SARS-CoV-2 test (CTM 2)

The 6800/8800 SARS-CoV-2 test is an automated real-time RT-PCR assay intended for the qualitative detection of nucleic acids from SARS-CoV-2 in healthcare provider-instructed, self-collected anterior nasal (nasal) swab samples (collected onsite) and healthcare provider-collected nasal, nasopharyngeal, and oropharyngeal swab samples collected from any individuals, including those suspected of COVID-19 by their healthcare provider, and those without symptoms or with other reasons to suspect COVID-19(28). The positive SARS-CoV-2 result is defined as positive on the SARS-CoV-2 channel (Target 1) and/or the pan-sarbeco channel (Target 2).

#### Hologic Aptima SARS-CoV-2 assay (tiebreaker; CTM 3)

As a commonly used molecular assay(30), CTM 3 was utilized as a tiebreaker. The test utilizes transcription-mediated amplification for the qualitative detection of RNA from SARS-CoV-2 isolated and purified from nasopharyngeal, nasal, mid-turbinate and oropharyngeal swab samples, nasopharyngeal wash/aspirate, or nasal aspirates obtained from individuals meeting COVID-19 clinical and/or epidemiological criteria(35).

### Analyses

All data analyses were performed using SAS/STAT^®^ software (v9.4 or higher of the SAS System for Linux). No formal sample size calculations were calculated for this study, but patient enrollment was adjusted to accommodate disease prevalence and to generate a minimum of 50 SARS-CoV-2 positives, 30 influenza A positives, and 10 influenza B positives for analysis.

The clinical performance of the POC SARS-CoV-2 & Influenza A/B test was evaluated using estimates of positive percent agreement (PPA), negative percent agreement (NPA), and overall percent agreement (OPA), calculated with 2-sided 95% confidence intervals (CI) using the Wilson-score method(36). Comparison was against either the composite comparator for SARS-CoV-2 (CTM 1, CTM 2, and CTM 3 [tiebreaker if required]) or a single comparator for influenza A/B (CTM 1).

For SARS-CoV-2, concordant results from CTM 1 and CTM 2 established true positive or true negative status. In the event of discordance between CTM 1 and CTM 2, CTM 3 (the tiebreaker) was used to establish true status. Samples could be coded as uninterpretable in the event of invalid, failed, aborted or missing results that were not resolved upon retesting. If CTM 1 or CTM 2 or CTM 3 returned uninterpretable results, and the other two methods returned different results (i.e., CTM 1 positive, CTM 2 negative, and CTM 3 uninterpretable), then the status of the sample would be described as indeterminate.

McNemar’s mid-p test (P-value) was used to assess any differences between sample types. A P-value of <0.05 was considered statistically significant.

POCT operators were asked to complete a questionnaire to evaluate the ease of use of the POC SARS-CoV-2 & Influenza A/B test.

*In silico* analysis of the POC SARS-CoV-2 & Influenza A/B test was performed to assess assay design inclusivity on all available SARS-CoV-2 sequences (taxonomy ID 2697049) in the Global Initiative on Sharing All Influenza Data (GISAID) and National Center for Biotechnology Information (NCBI) databases up to July 2024 (>16,800,000 sequences in NCBI and >8,900,000 sequences in GISAID). The predicted impact of each variant on gene target 1 and gene target 2 primer and probe binding site sequence was evaluated using Roche proprietary software and Melting5 software. A SARS-CoV-2 sequence would be potentially not detected by the POC SARS-CoV-2 & Influenza A/B test if a delay in cycle threshold greater than five cycles and/or a probe melting temperature of <65°C was predicted.

### Ethics statement

This study was a non-interventional evaluation of an *in vitro* diagnostic device (test results were not used to inform patient care decisions). This study was conducted in compliance with International Conference on Harmonisation Good Clinical Practice guidelines, regulations of the US Food and Drug Administration (FDA), and the Declaration of Helsinki. The study protocol was submitted to Institutional Review Boards (IRBs) in accordance with FDA and local regulatory requirements before the start of the study. The following IRBs gave permission for this study to be performed: NorthShore University Health System Institutional Review Board; University of Rochester, Research Subjects Review Board; and Western IRB.

## Results

### Participants and summary of testing results

Subjects from 10 geographically diverse healthcare facilities were enrolled in the study. The summary of testing results can be seen in **Supplemental Table A**. A total of 2,209 SARS-CoV-2 results from prospective samples (both nasal and nasopharyngeal swabs) were valid for inclusion in downstream analyses.

### Population and baseline characteristics

Baseline demographic data for study participants were available for the prospectively collected samples; details on the study participants are presented in **Table 1**. Of these subjects, 506 were male (47.8%) and 553 were female (52.2%). The median age was 35 years (range 0–86).

**Table 1:**
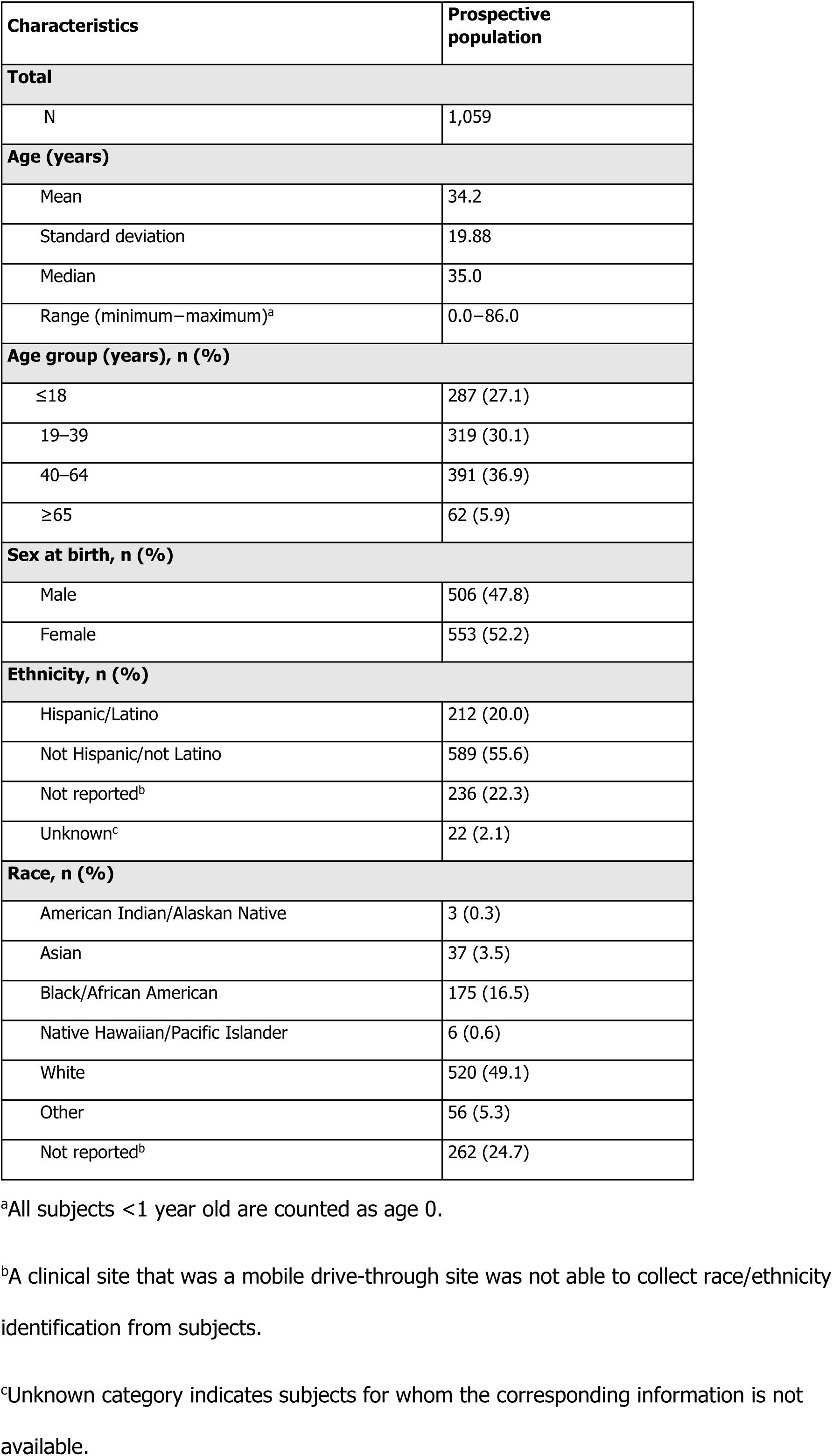
Demographics (prospective symptomatic and asymptomatic subjects)

Patient characteristics are present in **Supplemental Table B**. In total, 60.4% of subjects in the prospective population had signs and symptoms of respiratory infection, with days from onset of first symptom ranging from 1 to 365 days. The remaining 39.6% of subjects were asymptomatic but were clinically suspected of SARS-CoV-2 infection by their healthcare provider due to recent exposure or other reason. The nasal swab samples were evenly distributed between HCW-collected and self-collected.

### Medical history

The medical history relating to vaccination status is presented in **Table 2**. Most participants received a COVID-19 vaccine (68.7%), with Pfizer as the most common vaccine for both first and second doses (69.5% and 72.2%, respectively).

**Table 2:**
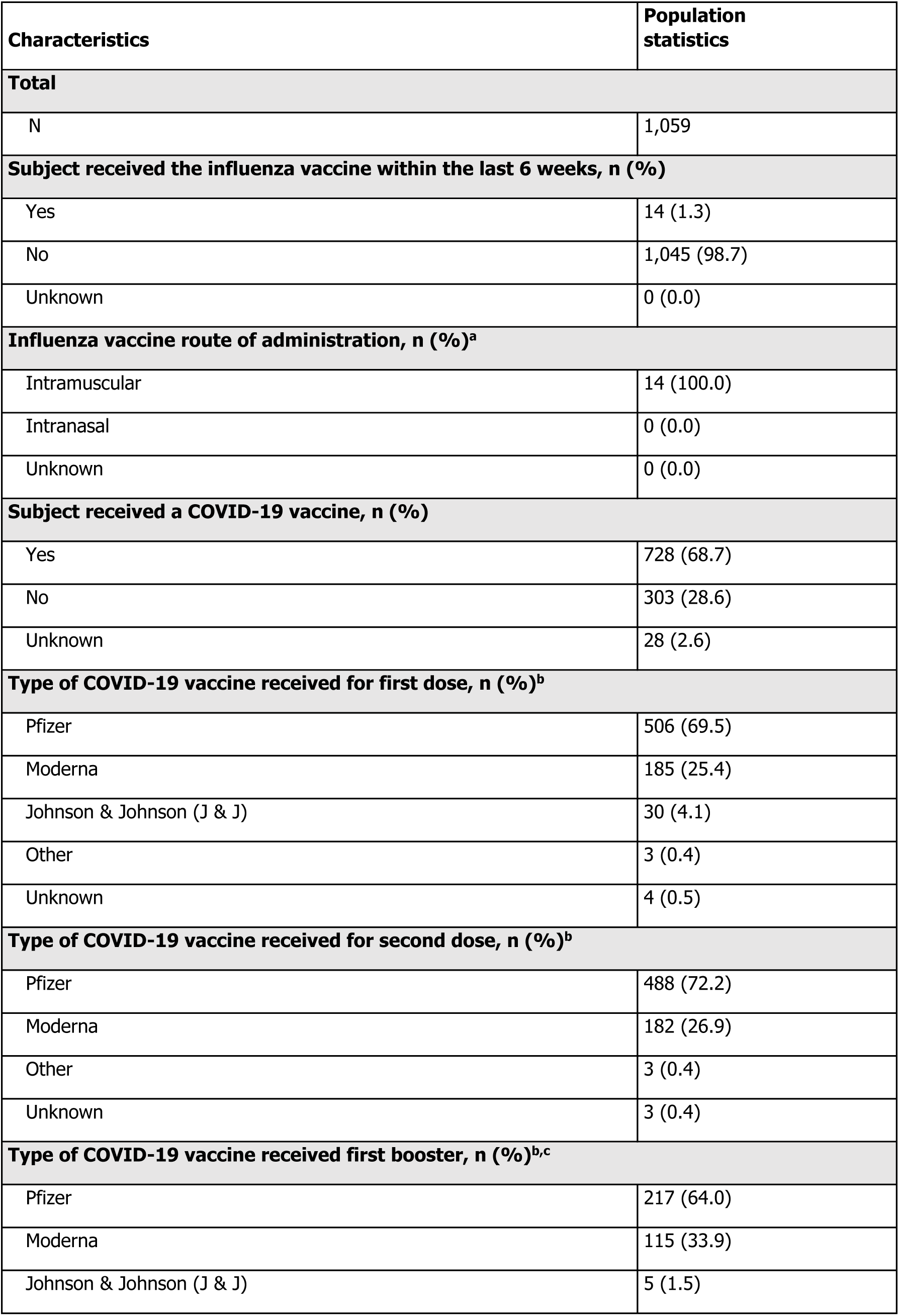
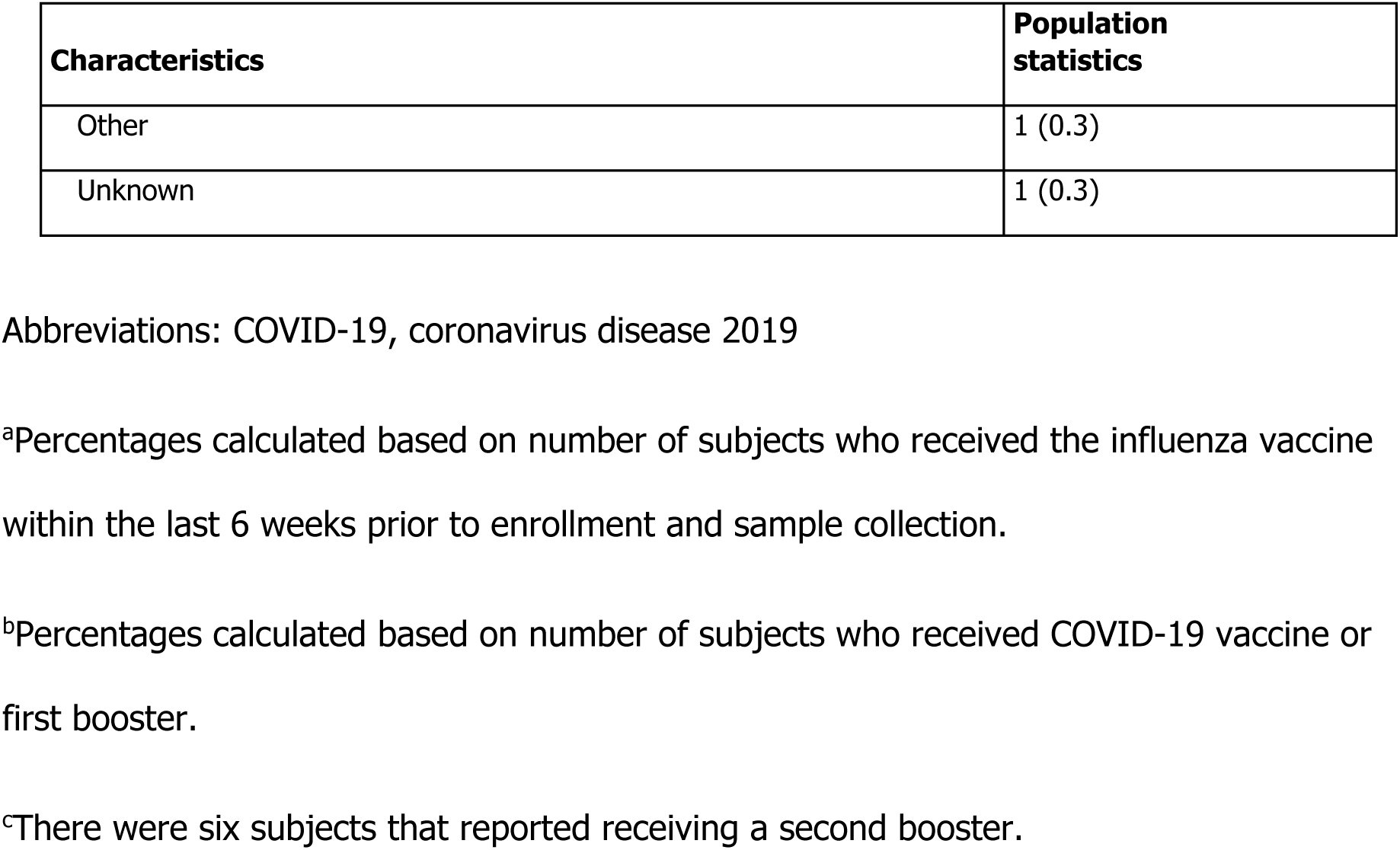
Subject medical history (prospective symptomatic and asymptomatic subjects)

### SARS-CoV-2

#### Determination of the composite comparator status for SARS-CoV-2

In symptomatic study participants, the composite comparator method was SARS-CoV-2 positive for 134 nasal and 131 nasopharyngeal swab samples (**Supplemental Table C**). In asymptomatic study participants, the composite comparator method was SARS-CoV-2 positive for 39 nasal and 39 nasopharyngeal swab samples (**Supplemental Table C**).

#### Performance

The total number of samples that were positive, negative, or invalid, and how these align with the composite comparator status, can be seen in **Figure 1**. The diagnostic performance (total number of positive and negative results) for SARS-CoV-2 can be seen in **Table 3**. Agreement between the composite comparator and the POC SARS-CoV-2 & Influenza A/B test for detection of SARS-CoV-2 in nasal swabs (total, prospective and retrospective, self-collected or HCW-collected), nasopharyngeal swabs (total, prospective and retrospective), in samples from vaccinated and unvaccinated participants, and in symptomatic and asymptomatic participants can be seen in **Supplemental Table D**. For nasal swab samples, the total OPA, PPA, and NPA was 98.8% (95% CI: 97.9, 99.3), 97.1% (95% CI: 93.3, 98.7), and 99.1% (95% CI: 98.2, 99.5), respectively. Of note, the OPA for HCW-collected nasal swab samples was 98.4% (95% CI: 97.0, 99.2) and 99.1% (95% CI: 98.0, 99.6) for self-collected nasal swab samples. The total OPA, PPA, and NPA for nasopharyngeal swab samples was 99.0% (95% CI: 98.2, 99.4), 96.4% (95% CI: 92.4, 98.4), and 99.4% (95% CI: 98.7, 99.8), respectively.

**Figure 1:**
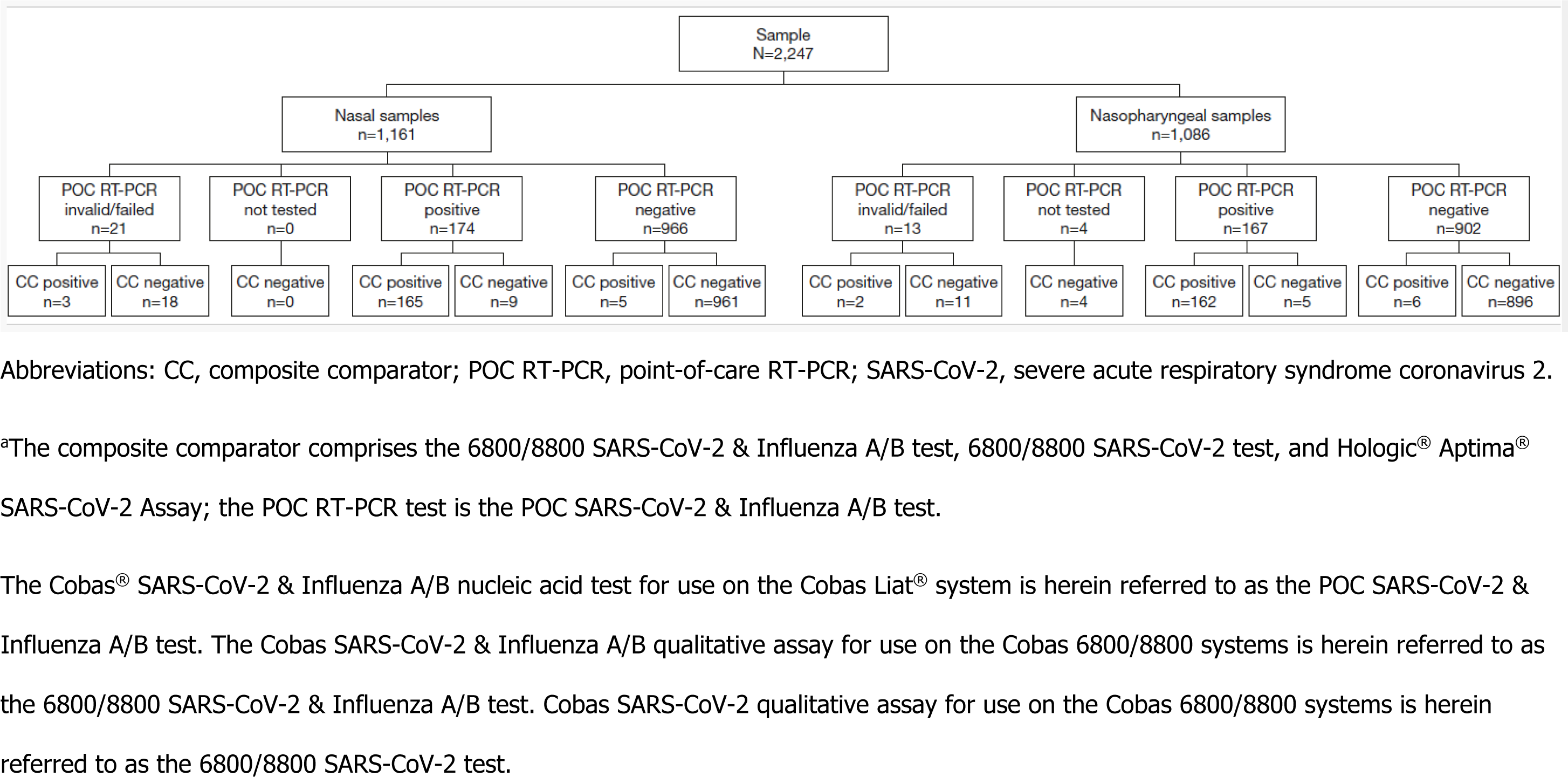
Detection of SARS-CoV-2 using the POC SARS-CoV-2 & Influenza A/B test and the composite comparator (prospective and retrospective, symptomatic and asymptomatic subjects)^a^

**Table 3:** Results for the detection of SARS-CoV-2 using the composite comparator compared with the POC SARS-CoV-2 & Influenza A/B test^a^.

|  | **Composite comparator (+)** | **Composite comparator (-)** | **Total** | **PPA % (95% CI)** | **NPA % (95% CI)** |
| --- | --- | --- | --- | --- | --- |
| **Nasal** |  |  |  |  |  |
| POC SARS-CoV-2 & Influenza A/B test (+) | 165 | 9 | 174 | **97.1 (93.3, 98.7)** | **99.1 (98.2, 99.5)** |
| POC SARS-CoV-2 & Influenza A/B test (-) | 5 | 961 | 966 |  |  |
| **Total** | **170** | **970** | **1,140** |  |  |
| **Nasopharyngeal** |  |  |  |  |  |
| POC SARS-CoV-2 & Influenza A/B test (+) | 162 | 5 | 167 | **96.4 (92.4, 98.4)** | **99.4 (98.7, 99.8)** |
| POC SARS-CoV-2 & Influenza A/B test (-) | 6 | 896 | 902 |  |  |
| **Total** | **168** | **901** | **1,069** |  |  |

Abbreviations: CI, Score confidence interval; NPA, negative percent agreement; PPA, positive percent agreement; SARS-CoV-2, severe acute respiratory syndrome coronavirus 2.
<sup>a</sup>The Cobas® SARS-CoV-2 & Influenza A/B nucleic acid test for use on the Cobas Liat® system is herein referred to as the POC SARS-CoV-2 & Influenza A/B test.

Using all paired results for nasal and nasopharyngeal swab samples by subject, the swab type did not make a difference in the reported result (McNemar’s mid-p test, P=0.771).

#### Cycle threshold (Ct) value distributions

The range of Ct values in nasal and nasopharyngeal swab samples can be seen in **Figure 2**. Self-collected nasal swabs showed the highest average viral load followed by nasopharyngeal swabs, then HCW-collected nasal swabs.

**Figure 2:**
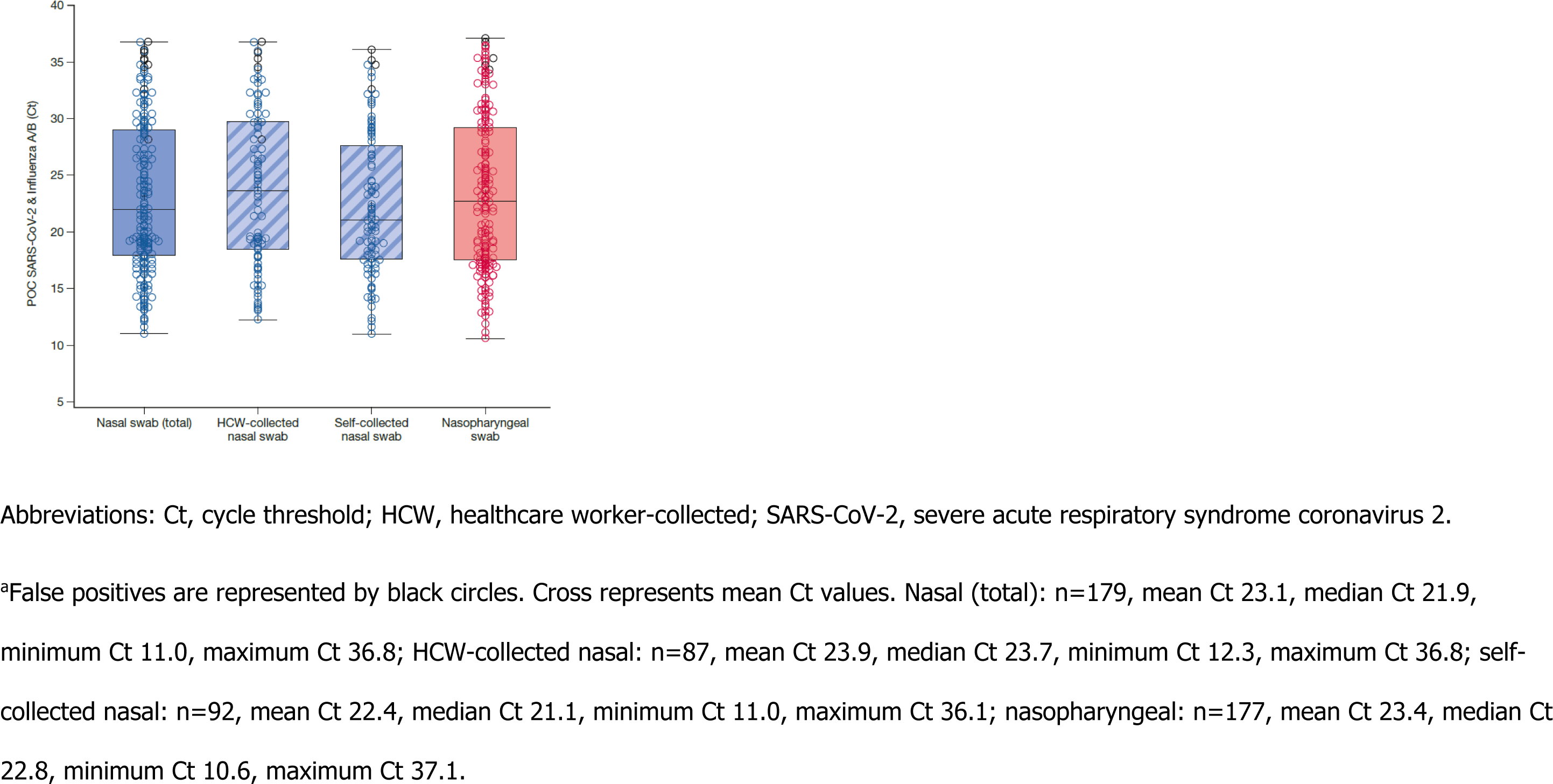
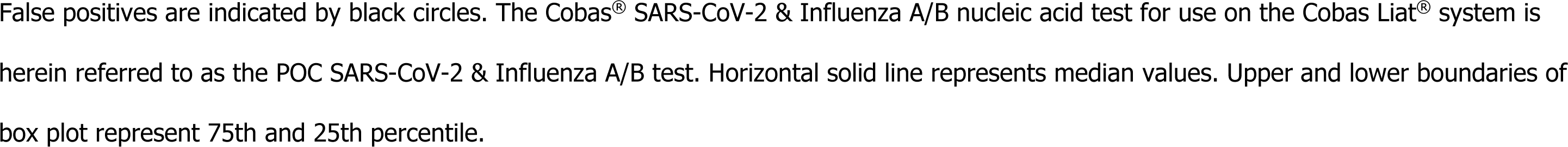
C_t_ values for the SARS-CoV-2 analyte from the POC SARS-CoV-2 & Influenza A/B test in nasal and nasopharyngeal swab samples^a^

The range of Ct values in subject-paired nasal and nasopharyngeal swab samples can be seen in **Figure 3**. The Deming regression analysis showed that the two sample types were highly correlated (**Figure 3A**; r = 0.83). Exploration of the collection method showed that HCW-collected nasal swabs had greater concordance between the paired samples (**Figure 3B**; r = 0.91) than the self-collected nasal swabs (r = 0.77; **Figure 3C**).

**Figure 3:**
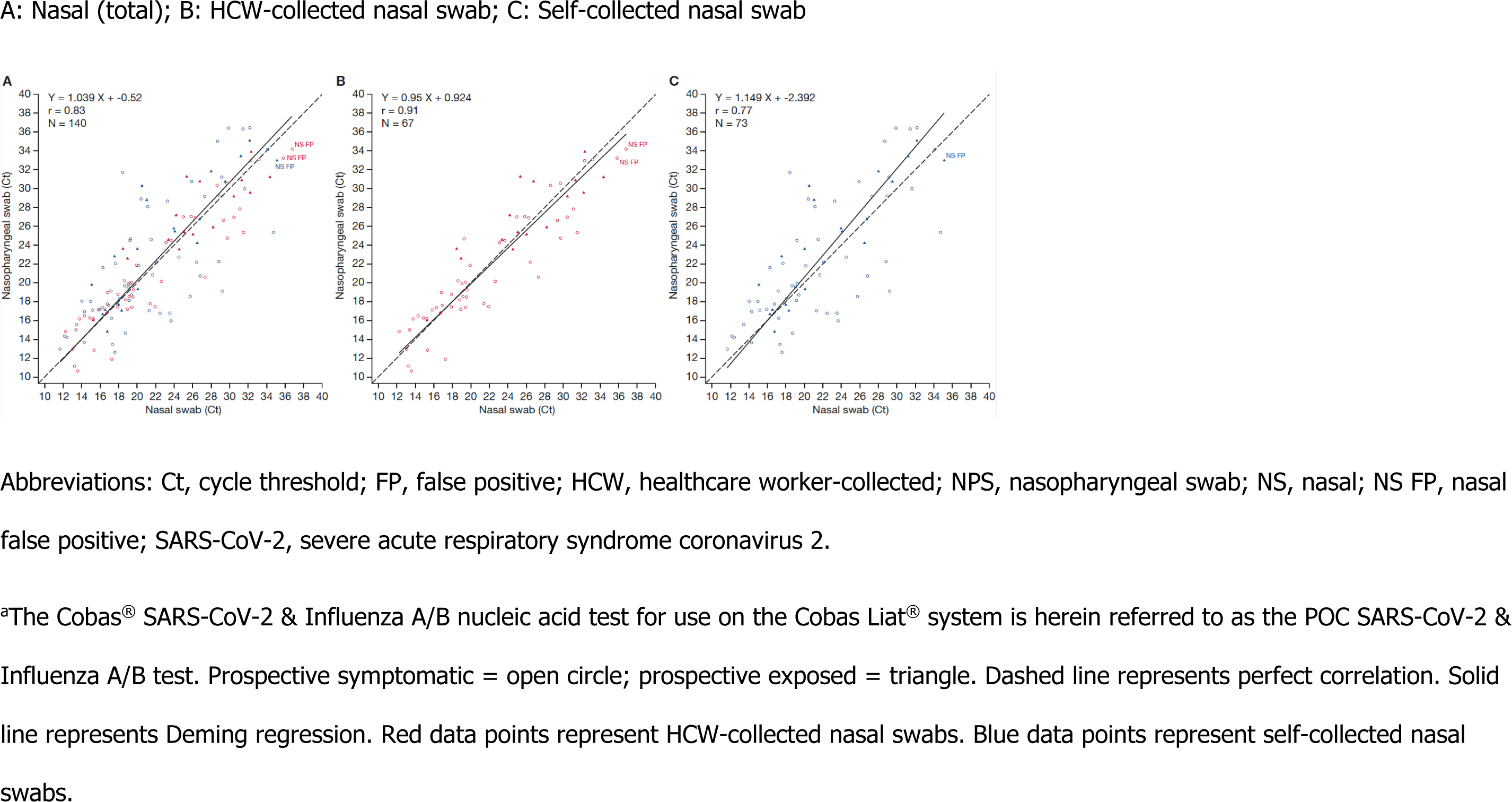
Deming regression for the SARS-CoV-2 analyte from the POC SARS-CoV-2 & Influenza A/B test in subject-paired nasal and nasopharyngeal swab samples^a^

#### Discordance

Fourteen nasal and 11 nasopharyngeal swab samples showed discordant results between the POC SARS-CoV-2 & Influenza A/B test and the centralized testing methods (**Supplemental Table E**); 10 were positive only on the POC SARS-CoV-2 & Influenza A/B test.

### Influenza A

#### Performance

The agreement between the POC SARS-CoV-2 & Influenza A/B test and the 6800/8800 SARS-CoV-2 & Influenza A/B test for detection of influenza A in nasopharyngeal swabs (total, prospective and retrospective) and nasal swabs (total, prospective and retrospective, HCW-collected or self-collected) can be seen in **Supplemental Table F**. The OPA ranged from 98.9% to 100.0%.

#### Discordance

Two nasal and five nasopharyngeal swab samples showed discordant results between the POC SARS-CoV-2 & Influenza A/B test and the 6800/8800 SARS-CoV-2 & Influenza A/B test for detection of influenza A (**Supplemental Table G**), four of which were positive only on the POC SARS-CoV-2 & Influenza A/B test.

### Influenza B

#### Performance

The agreement between the POC SARS-CoV-2 & Influenza A/B test and the 6800/8800 SARS-CoV-2 & Influenza A/B test for detection of influenza B in nasopharyngeal swabs (total, prospective and retrospective) and nasal swabs (total, prospective and retrospective, self-collected nasal or HCW-collected) can be seen in **Supplemental Table H**. The OPA for all comparisons was 100.0%.

#### Discordance

No discordant results between the POC SARS-CoV-2 & Influenza A/B test and the 6800/8800 SARS-CoV-2 & Influenza A/B test for detection of influenza B were recorded.

### Evaluation of the ease of use for the Cobas Liat system

Twenty-seven operators with experience of testing samples on the Cobas Liat completed the questionnaire.The operators’ average scores indicating their agreement with the statements in the questionnaire are shown in **Supplemental Table I**. The overall score was 4.5 out of 5 for the operators’ answers to all eight statements, indicating that the operators agreed that the Cobas Liat system was easy to use.

### Inclusivity analysis

The POC SARS-CoV-2 & Influenza A/B test targets the nucleocapsid (N) and ORF1a/b regions of the SARS-CoV-2 genome.(34) *In silico* analysis showed that 99.98% of NCBI and 99.99% of GISAID sequences for SARS-CoV-2 had no changes in the primer/probe binding sites of both target regions simultaneously. All sequences were predicted to be detected by at least one of the two sites.

## Discussion

In this study, we evaluated the performance of the POC SARS-CoV-2 & Influenza A/B test against up to three centralized assays, stratified by various parameters. The high performance of the POC SARS-CoV-2 & Influenza A/B test in comparison with centralized testing methods utilizing nasopharyngeal or nasal samples has previously been reported(37–41), but this study is the first to report the performance of the POC SARS-CoV-2 & Influenza A/B test across variables such as collection method (self/HCW, retrospective/prospective), vaccination status, symptom status, and ease of use. Using the comparator result to establish the status of infection, we found high agreement between the POC RT-PCR test and the centralized testing assays for all three analytes across all variables.

Previously, a meta-analysis of nucleic acid amplification testing reported that the sensitivity for detection of SARS-CoV-2 in nasal samples may be less than nasopharyngeal samples(42), whilst a study specifically assessing the POC SARS-CoV-2 & Influenza A/B test reported similar differences in SARS-CoV-2 detection by sample type(37). However, we saw no evidence of a sample-type difference in sensitivity, with SARS-CoV-2 positivity identified in both nasal and nasopharyngeal swab samples to a similar extent (POC SARS-CoV-2 & Influenza A/B test positivity rate approximately 15%; the difference in sensitivity [nasal minus nasopharyngeal] was +0.9%).

A small study from Denmark found almost equivalent sensitivity for detection of SARS-CoV-2 using self- and HCW-collected samples (84.2% vs 89.5%, respectively), and that patients preferred self-collection(43). In our study, whether nasal samples were collected by HCWs or by the patients themselves had little impact on the agreement values between the POC RT-PCR test and the comparators. Indeed, in all comparisons the agreement for self-collected samples was actually higher than for HCW-collected samples. For SARS-CoV-2, Ct values of paired nasal and nasopharyngeal swab samples as measured by the POC RT-PCR test were highly correlated, and Ct values were on average lower in nasal swab samples (i.e., higher viral RNA titer) compared with nasopharyngeal swab samples, with self-collected nasal swabs exhibiting the lowest Ct values. Collectively, this indicates that the high performance level of the POC RT-PCR test is maintained across collection methods, and that patients are able to self-sample effectively, and are willing to do so.

We identified a small number of discrepant samples for the SARS-CoV-2 analyte in both nasal and nasopharyngeal swab samples, consistent with other studies(37, 38), most of which were false positives. The majority of the false positives were in the high Ct/low viral titer range (POC SARS-CoV-2 & Influenza A/B test Ct values: lowest 28.2, median 35.2, highest 37.1) and thus close to or at the lower limit of detection of Ct 35.2 (USA-WA1/2020 strain) for the POC SARS-CoV-2 & Influenza A/B test(34). Of the false negatives detected by the POC SARS-CoV-2 & Influenza A/B test, all except one sample were in the high Ct range. Indeed, after exploratory re-testing of false-negative nasopharyngeal samples, two were found to be positive (re-tested Ct values were 34.8 and 35.4). Generally, low viral burden can be a cause of discrepant results(44), and a small external quality assessment (reproducibility) study reported that the performance of the POC SARS-CoV-2 & Influenza A/B test is highest at low Ct values(45).

US legislation categorizes tests for complexity (moderate or high) using seven criteria, such as the need for training and experience, and assigns scores within each criterion(45). Test systems can be assigned as ‘waived complexity’ under certain conditions, and these tests require no formal operator training or competency(45). In addition to CLIA-certified laboratories, the POC SARS-CoV-2 & Influenza A/B test is authorized for use in patient care settings that are CLIA waived(34). In conjunction with the fast turnaround offered by the Cobas Liat system and demonstrated in studies of the POC SARS-CoV-2 & Influenza A/B test in real-world settings (40, 46) or studies evaluating workflow or processing time(41, 47), our study confirms that the POC SARS-CoV-2 & Influenza A/B test is easy to use in CLIA-waived settings, and offers results in a timeframe conducive for rapid patient management.

The strength of our multicenter study lies in the comprehensive nature of the variables assessed, encompassing a wide variety of patient characteristics likely to be encountered in healthcare settings. The study included a prospectively enrolled cohort of patients seeking care, which is representative of the intended-use population of the POC SARS-CoV-2 & Influenza A/B test. Our study has some limitations. Firstly, we did not ascertain the SARS-CoV-2 variants present in the samples. Whilst the emergence of SARS-CoV-2 variants has the ability to affect the diagnostic performance of the test, previous studies have indicated that the POC SARS-CoV-2 & Influenza A/B test detects both wild-type and variants of concern, such as Alpha (B.1.1.7) or Omicron (B.1.1.529)(39, 47). Bioinformatic analysis to assess inclusivity showed that the POC SARS-CoV-2 & Influenza A/B test is predicted to bind all sequences available in the NCBI and GISAID databases as of July 2024. The dual target assay design helps to ensure that the assay is robust and safeguards against the emergence of variants that have the potential to affect assay performance and evade detection.

## Conclusion

A POC RT-PCR test combining both measurement of SARS-CoV-2 and influenza subtypes with performance equivalent to routine centralized testing would provide a critical tool to improve the diagnosis and management of COVID-19 and influenza. We found that the performance of the POC SARS-CoV-2 & Influenza A/B test was comparable to centralized testing methods. With the ease of use and equivalent performance, it highlights the question as to whether centralized testing should be considered as the gold standard. A study of the POC Cobas Influenza A/B test (a Liat RT-PCR test that detects only the influenza analyte) in the emergency department found that POC testing for influenza was useful in improving several metrics, including the indication for treatment with neuraminidase in positive cases(48). It is relevant to note that SARS-CoV-2 and influenza coinfection can increase the risk of severe outcomes compared with those infected with SARS-CoV-2 alone, particularly for those coinfected with influenza A.(49) Our study highlights the benefits of molecular multiplex POCT to help improve timely differentiation of respiratory diseases that share similar symptoms and support efforts to improve patient management.

## Assay disclaimers

The Cobas SARS-CoV-2 & Influenza A/B qualitative assay for use on the Cobas Liat system (herein referred to as the POC SARS-CoV-2 & Influenza A/B test) was originally approved under Emergency Use Authorization EUA201779 and has since been cleared and Clinical Laboratory Improvement Amendments (CLIA) waived under 223591/CW220014, respectively, in the US and is CE-IVD marked in the European Union. Sample collection in the patient’s home is not approved in the US or European Union.

The Cobas SARS-CoV-2 & Influenza A/B qualitative assay for use on the Cobas 6800/8800 systems (herein referred to as the 6800/8800 SARS-CoV-2 & Influenza A/B test) is authorized only for use under Emergency Use Authorization (EUA) in the US, and is CE-IVD marked in the European Union. The assay is not approved for use in asymptomatic patients in the US or European Union. The assay is not approved for use as a point-of-care (POC) test/near-patient test (NPT) in the US/European Union. Sample collection in the patient’s home is not approved in the US or European Union.

The Cobas SARS-CoV-2 qualitative assay for use on the Cobas 6800/8800 systems (herein referred to as the 6800/8800 SARS-CoV-2 test) is authorized only for use under EUA in the US, and is CE-IVD marked in the European Union. The assay is not approved for use as a POC test/NPT in the US/European Union. Sample collection in the patient’s home is not approved in the US or European Union.

Hologic Aptima SARS-CoV-2 assay is an FDA EUA assay.(30)

## Data availability

The datasets generated during and/or analyzed during the current study are not publicly available due to patient confidentiality. Any access requests from qualified researchers should be submitted directly to the Ethical Committee of each participating study site.

## Acknowledgments

Medical writing support was provided by Corrinne Segal of Obsidian Healthcare Group Ltd (London, UK) and was funded by Roche Molecular Systems, Inc.

COBAS and LIAT are trademarks of Roche. All other product names and trademarks are the property of their respective owners.

We would like to thank Saima Shams and Vaishali Mody for their contributions to the development of the Cobas SARS-CoV-2 & Influenza A/B qualitative assay for use on the Cobas Liat system (herein referred to as the POC SARS-CoV-2 & Influenza A/B test), and Jingtao Sun for his bioinformatics expertise.

