## Supplemental for "A multicenter study to assess the performance of the point-of-care RT-PCR Cobas SARS-CoV-2 & Influenza A/B nucleic acid test for use on the Cobas Liat system in comparison with centralized assays across healthcare facilities in the United States"

### Title

Elissa M. Robbins^a#^

Rasa Bertuzis^a^

Ho-Chen Chiu^b^

Lupe Miller^a^

Christopher Noutsios^a^

### Affiliations

^a^Roche Molecular Systems, Inc., Pleasanton, California, USA

^b^Roche Sequencing Solutions, Inc., Indianapolis, Indiana, USA

### ^#^Corresponding author

Elissa M. Robbins

### Running title

POC SARS-CoV-2 & Influenza A/B test

### Supplementary material

###### Signs/symptoms of suspected respiratory viral infection consistent with COVID-19

- Fever or chills
- Cough
- Shortness of breath or difficulty breathing
- Sore throat
- Congestion or runny nose
- Nausea or vomiting
- Muscle or body aches
- Headaches
- Fatigue
- Diarrhea
- Loss of taste or smell
- Other, specify______

###### Eligibility criteria

Inclusion

- Adult or pediatric, as long as provided written informed consent (per IRB)

Exclusion

- Prior enrollment in this study
- Both nostrils already used for same-day SOC specimen generation (nasal sampling)
- Both nostrils already used for same-day SOC specimen generation (nasopharyngeal sampling)
- Except for SOC sampling (if collected), same-day nasal or nasopharyngeal cavity sampling
- Nasopharyngeal or nasal aspirate and nasopharyngeal or nasal wash performed same day
- Contraindication to nasopharyngeal cavity sampling as performed according to the healthcare facility’s site policies and procedures
- Receipt of any formulation of antiviral medication in the preceding 7 days, including but not limited to rimantadine (Flumadine), amantadine (Gocovri), oseltamivir (Tamiflu), zanamivir (Relenza), peramivir (Rapivab), baloxavir marboxil (Xofluza), ribavirin (Copegus, Rebetol, Ribasphere, and Virazole), and remdesivir (GS-5734)
- Receipt of topical mupirocin in the preceding 7 days
- Receipt of influenza vaccination that was administered through the nasopharynx (e.g., FluMist) within the preceding 6 weeks

Abbreviations: COVID-19, coronavirus disease 2019; IRB, institutional review board; SOC, standard of care

###### Supplemental Table A: Summary of the POC SARS-CoV-2 & Influenza A/B test results for the detection of SARS-CoV-2 stratified by sample or collection type^a^

|  | **Cobas Liat SARS-CoV-2 & Influenza A/B test** | **SARS-CoV-2 channel Total** | **Influenza A** **channel Total** | **Influenza B** **channel Total** |
| --- | --- | --- | --- | --- |
|  | **Nasal (HCW-collected and self)** | |  |  |
|  | Valid (positive and negative) | 1,140 | 1,208 | 1,208 |
|  | Invalid/failed | 21 | 22 | 22 |
|  | Not tested | 0 | 0 | 0 |
|  | Total | 1,161 | 1,230 | 1,230 |
|  | **Nasal HCW-collected** | |  |  |
|  | Valid (positive and negative) | 566 | 634 | 634 |
|  | Invalid/failed | 10 | 11 | 11 |
|  | Not tested | 0 | 0 | 0 |
|  | Total | 576 | 645 | 645 |
|  | **Nasal self-collected** | |  |  |
|  | Valid (positive and negative) | 574 | 574 | 574 |
|  | Invalid/failed | 11 | 11 | 11 |
|  | Not tested | 0 | 0 | 0 |
|  | Total | 585 | 585 | 585 |
| **Nasopharyngeal** | | |  |  |
| Valid (positive and negative) | | 1,069 | 1,199 | 1,196 |
| Invalid/failed | | 13 | 13 | 13 |
| Not tested | | 4 | 4 | 7 |
| Total | | 1,086 | 1,216 | 1,216 |
|  | **Total** | |  |  |
|  | Valid (positive and negative) | 2,209 | 2,407 | 2,404 |
|  | Invalid/failed | 34 | 35 | 35 |
|  | Not tested | 4 | 4 | 7 |
|  | Total | 2,247 | 2,446 | 2,446 |

Abbreviations: HCW, healthcare-worker; SARS-CoV-2, severe acute respiratory syndrome coronavirus 2.

^a^The Cobas^®^ SARS-CoV-2 & Influenza A/B nucleic acid test for use on the Cobas Liat^®^ system is herein referred to as the POC SARS-CoV-2 & Influenza A/B test.

###### Supplemental Table B: Patient characteristics (prospective symptomatic and asymptomatic subjects)

| **Characteristics** | **Population statistics** |
| --- | --- |
| **Total** |  |
| N | 1,059 |
| **Patient status at the time of consent, n (%)** | |
| Inpatient | 13 (1.2) |
| Outpatient | 1046 (98.8) |
| Other | 0 (0.0) |
| **Reason for COVID-19 testing, n (%)** | |
| Symptomatic | 640 (60.4) |
| Asymptomatic: suspected due to recent exposure or other reason | 419 (39.6) |
| **Signs and symptoms, n (%)** |  |
| Yes | 640 (60.4) |
| No | 419 (39.6) |
| **Days from onset of first symptom** | |
| N^a^ | 629 |
| Mean | 5.7 |
| Standard deviation | 22.20 |
| Median | 3.0 |
| Range (minimum−maximum) | 1−365 |
| **Symptoms, n (%)** | |
| Congestion or runny nose | 342 (53.4) |
| Cough | 304 (47.5) |
| Sore throat | 258 (40.3) |
| Headaches | 189 (29.5) |
| Fever or chills | 184 (28.8) |
| Fatigue | 138 (21.6) |
| Muscle or body aches | 116 (18.1) |
| Nausea or vomiting | 87 (13.6) |
| Shortness of breath or difficulty breathing | 61 (9.5) |
| Diarrhea | 40 (6.3) |
| Other^b^ | 37 (5.8) |
| Loss of taste or smell | 10 (1.6) |
| **Nasal collection method, n (%)** | |
| HCW-collected | 530 (50.0) |
| Self-collected | 529 (50.0) |
| Unknown | 0 (0.0) |

Abbreviations: COVID-19, coronavirus disease 2019; HCW, healthcare-worker.

^a^11 of the 640 subjects that reported being symptomatic were missing days from onset of first symptoms.

^b^Other includes: cold symptoms, allergies, bloody nose, cold, chest tightness during cough, dehydrated, dizziness, ear pain, sinus pressure, earache, hoarseness, itchy eyes, lightheaded, myalgia, nasal drainage, sinus infection, sneezing, stomach pain, scratchy throat, sinus issues, swollen lymph nodes, teary eyes and sneezing, tickle in throat, watery eyes, dizziness, poor appetite, sneezing, stomach ache.

###### Supplemental Table C: Determination of composite comparator status for SARS-CoV-2 (prospective and retrospective, symptomatic and asymptomatic subjects)^a^

|  | **6800/8800 SARS-CoV-2 & Influenza A/B test (CTM 1)** | **6800/8800 SARS-CoV-2 test (CTM 2)** | **Hologic^®^ Aptima^®^ SARS-CoV-2 Assay**  **(CTM 3)** | **Composite comparator interpretation** | **Counts** |
| --- | --- | --- | --- | --- | --- |
| **Symptomatic** | | | | | |
| Nasal (n=747) | | | | | |
|  | + | + | NR | + | 133 |
|  | + | - | - | - | 1 |
|  | - | - | NR | - | 612 |
|  | - | + | + | + | 1 |
| Nasopharyngeal (n=673) | | | | | |
|  | + | + | NR | + | 131 |
|  | + | - | - | - | 3 |
|  | - | - | NR | - | 536 |
|  | - | + | - | - | 3 |
| **Asymptomatic** | | | | | |
| Nasal (n=414) | | | | | |
|  | + | + | NR | + | 39 |
|  | + | **-** | **-** | - | 3 |
|  | - | - | NR | - | 371 |
|  | - | + | - | - | 1 |
| Nasopharyngeal (n=413) | | | | | |
|  | + | + | NR | + | 39 |
|  | + | - | - | - | 3 |
|  | - | - | NR | - | 369 |
|  | - | + | - | - | 2 |

Abbreviations: CTM, centralized testing method; NR, not required (when the two comparator test results are concordant); +, positive test result; -, negative test result; SARS-CoV-2, severe acute respiratory syndrome coronavirus 2.

^a^The Cobas^®^ SARS-CoV-2 & Influenza A/B qualitative assay for use on the Cobas 6800/8800 systems is herein referred to as the 6800/8800 SARS-CoV-2 & Influenza A/B test. Cobas SARS-CoV-2 qualitative assay for use on the Cobas 6800/8800 systems is herein referred to as the 6800/8800 SARS-CoV-2 test.

###### Supplemental Table D: Diagnostic performance for the detection of SARS-CoV-2 using composite comparator compared with the POC SARS-CoV-2 & Influenza A/B test (prospective and retrospective, symptomatic and asymptomatic subjects)^a^

|  | **Composite comparator (+) and POC SARS-CoV-2 & Influenza A/B test (+)** | **Composite comparator (+) and POC SARS-CoV-2 & Influenza A/B test (-)** | **Composite comparator (-) and POC SARS-CoV-2 & Influenza A/B test (+)** | **Composite comparator (-) and POC SARS-CoV-2 & Influenza A/B test (-)** | **OPA %**  **(95% CI)** | **PPA %**  **(95% CI)** | **NPA %**  **(95% CI)** |
| --- | --- | --- | --- | --- | --- | --- | --- |
| **Total (nasal and nasopharyngeal)** | | | | | | | |
| ***Total (n=2,209)*** | ***327*** | ***11*** | ***14*** | ***1,857*** | ***98.9***  ***(98.3, 99.2)*** | ***96.8***  ***(94.3, 98.2)*** | ***99.3***  ***(98.7, 99.6)*** |
| Symptomatic (n=1,396) | 252 | 9 | 10 | 1,125 | 98.6  (97.9, 99.1) | 96.6  (93.6, 98.2) | 99.1  (98.4, 99.5) |
| Asymptomatic (n=813) | 75 | 2 | 4 | 732 | 99.3  (98.4, 99.7) | 97.4  (91.0, 99.3) | 99.5  (98.6, 99.8) |
| **Nasal (symptomatic and asymptomatic)** | | | | | | | |
| ***Total (n=1,140)*** | ***165*** | ***5*** | ***9*** | ***961*** | ***98.8***  ***(97.9, 99.3)*** | ***97.1***  ***(93.3, 98.7)*** | ***99.1***  ***(98.2, 99.5)*** |
| Prospective sampling (n=1,022) | 142 | 5 | 6 | 869 | 98.9  (98.1, 99.4) | 96.6  (92.3, 98.5) | 99.3  (98.5, 99.7) |
| Retrospective sampling (n=118) | 23 | 0 | 3 | 92 | 97.5  (92.8, 99.1) | 100.0  (85.7, 100.0) | 96.8  (91.1, 98.9) |
| HCW-collected (n=566) | 80 | 4 | 5 | 477 | 98.4  (97.0, 99.2) | 95.2  (88.4, 98.1) | 99.0  (97.6, 99.6) |
| Self-collected (n=574) | 85 | 1 | 4 | 484 | 99.1  (98.0, 99.6) | 98.8  (93.7, 99.8) | 99.2  (97.9, 99.7) |
| **Nasopharyngeal (symptomatic and asymptomatic)** | | | | | | | |
| ***Total (n=1,069)*** | ***162*** | ***6*** | ***5*** | ***896*** | ***99.0***  ***(98.2, 99.4)*** | ***96.4***  ***(92.4, 98.4)*** | ***99.4***  ***(98.7, 99.8)*** |
| Prospective sampling (n=1,023) | 139 | 6 | 5 | 873 | 98.9  (98.1, 99.4) | 95.9  (91.3, 98.1) | 99.4  (98.7, 99.8) |
| Retrospective sampling (n=46) | 23 | 0 | 0 | 23 | 100.0  (92.3, 100.0) | 100.0  (85.7, 100.0) | 100.0  (85.7, 100.0) |
| **SARS-CoV-2 vaccination status^b^** | | | | | | | |
| **Vaccinated** | | | | | | | |
| ***Total (n=1,451)*** | ***230*** | ***8*** | ***8*** | ***1,205*** | ***98.9***  ***(98.2, 99.3)*** | ***96.6***  ***(93.5, 98.3)*** | ***99.3***  ***(98.7, 99.7)*** |
| Nasal (n=738) | 118 | 3 | 4 | 613 | 99.1  (98.1, 99.5) | 97.5  (93.0, 99.2) | 99.4  (98.3, 99.7) |
| HCW-collected (n=369) | 55 | 2 | 2 | 310 | 98.9  (97.2, 99.6) | 96.5  (88.1, 99.0) | 99.4  (97.7, 99.8) |
| Self-collected (n=369) | 63 | 1 | 2 | 303 | 99.2  (97.6, 99.7) | 98.4  (91.7, 99.7) | 99.3  (97.6, 99.8) |
| Nasopharyngeal (n=713) | 112 | 5 | 4 | 592 | 98.7  (97.6, 99.3) | 95.7  (90.4, 98.2) | 99.3  (98.3, 99.7) |
| **Unvaccinated** | | | | | | | |
| ***Total (n=702)*** | ***88*** | ***3*** | ***6*** | ***605*** | ***98.7***  ***(97.6, 99.3)*** | ***96.7***  ***(90.8, 98.9)*** | ***99.0***  ***(97.9, 99.5)*** |
| Nasal (n=374) | 43 | 2 | 5 | 324 | 98.1  (96.2, 99.1) | 95.6  (85.2, 98.8) | 98.5  (96.5, 99.3) |
| HCW-collected (n=187) | 25 | 2 | 3 | 157 | 97.3  (93.9, 98.9) | 92.6  (76.6, 97.9) | 98.1  (94.6, 99.4) |
| Self-collected (n=187) | 18 | 0 | 2 | 167 | 98.9  (96.2, 99.7) | 100.0  (82.4, 100.0) | 98.8  (95.8, 99.7) |
| Nasopharyngeal (n=328) | 45 | 1 | 1 | 281 | 99.4  (97.8, 99.8) | 97.8  (88.7, 99.6) | 99.6  (98.0, 99.9) |
| **Symptom status^c^** | | | | | | | |
| **Symptomatic** | | | | | | | |
| ***Total (n=1,396)*** | ***252*** | ***9*** | ***10*** | ***1,125*** | ***98.6***  ***(97.9, 99.1)*** | ***96.6***  ***(93.6, 98.2)*** | ***99.1***  ***(98.4, 99.5)*** |
| Nasal (n=734) | 128 | 4 | 7 | 595 | 98.5  (97.3, 99.2) | 97.0  (92.5, 98.8) | 98.8  (97.6, 99.4) |
| HCW-collected (n=367) | 62 | 3 | 4 | 298 | 98.1  (96.1, 99.1) | 95.4  (87.3, 98.4) | 98.7  (96.6, 99.5) |
| Self-collected (n=367) | 66 | 1 | 3 | 297 | 98.9  (97.2, 99.6) | 98.5  (92.0, 99.7) | 99.0  (97.1, 99.7) |
| Nasopharyngeal (n=662) | 124 | 5 | 3 | 530 | 98.8  (97.6, 99.4) | 96.1  (91.2, 98.3) | 99.4  (98.4, 99.8) |
| **Asymptomatic** | | | | | | | |
| ***Total (n=813)*** | ***75*** | ***2*** | ***4*** | ***732*** | ***99.3***  ***(98.4, 99.7)*** | ***97.4***  ***(91.0, 99.3)*** | ***99.5***  ***(98.6, 99.8)*** |
| Nasal (n=406) | 37 | 1 | 2 | 366 | 99.3  (97.9, 99.7) | 97.4  (86.5, 99.5) | 99.5  (98.0, 99.9) |
| HCW-collected (n=199) | 18 | 1 | 1 | 179 | 99.0  (96.4, 99.7) | 94.7  (75.4, 99.1) | 99.4  (96.9, 99.9) |
| Self-collected (n=207) | 19 | 0 | 1 | 187 | 99.5  (97.3, 99.9) | 100.0  (83.2, 100.0) | 99.5  (97.0, 99.9) |
| Nasopharyngeal (n=407) | 38 | 1 | 2 | 366 | 99.3  (97.9, 99.7) | 97.4  (86.8, 99.5) | 99.5  (98.0, 99.9) |

Abbreviations: CI, Score confidence interval; HCW, healthcare worker; NPA, negative percent agreement; OPA, overall percent agreement; PPA, positive percent agreement; SARS-CoV-2, severe acute respiratory syndrome coronavirus 2.

^a^The Cobas^®^ SARS-CoV-2 & Influenza A/B nucleic acid test for use on the Cobas Liat^®^ system is herein referred to as the POC SARS-CoV-2 & Influenza A/B test.

^b^Vaccination status for retrospective vendor-acquired samples was not available and therefore can not be categorized (n=46).

^c^Prospective samples.

###### Supplemental Table E: Discordant results for the detection of SARS-CoV-2 using composite comparator compared with the POC SARS-CoV-2 & Influenza A/B test (negative or positive)^a^

| **Population** | **POC RT-PCR**  **POC SARS-CoV-2 & Influenza A/B test**  **Status (Ct value)** | **CTM 1**  **6800/8800 SARS-CoV-2 & Influenza A/B test**  **Status (Ct value)** | **CTM 2**  **6800/8800 SARS-CoV-2 test**  **Status (Ct value)** | **CTM 3**  **Hologic Aptima SARS-CoV-2 Assay**  **Status (total) RLU (x1,000)** | **Composite comparator** | **Discordant type** |
| --- | --- | --- | --- | --- | --- | --- |
| **Nasal** | | | | | | |
| **Self-collected** | | | | | | |
| Asymptomatic | Positive (35.1) | Negative | Negative | NA | Negative | False positive |
| Symptomatic | Positive (32.6) | Negative | Negative | NA | Negative | False positive |
| Symptomatic | Positive (34.7) | Negative | Negative | NA | Negative | False positive |
| Symptomatic | Positive (36.1) | Negative | Negative | NA | Negative | False positive |
| Symptomatic | Negative | Negative | Positive (NaN/38.6) | Positive (753) | Positive | False negative |
| **HCW-collected** | | | | | | |
| Asymptomatic | Positive (34.5) | Negative | Negative | NA | Negative | False positive |
| Asymptomatic | Negative | Positive (26.5/25.9) | Positive (26.6/26.5) | NA | Positive | False negative^b^ |
| Symptomatic | Positive (35.3) | Negative | Negative | NA | Negative | False positive |
| Symptomatic | Positive (28.2) | Negative | Negative | NA | Negative | False positive |
| Symptomatic | Positive (35.8) | Positive (NaN/36.1) | Negative | Negative (286) | Negative | False positive |
| Symptomatic | Positive (36.8) | Negative | Negative | NA | Negative | False positive |
| Symptomatic | Negative | Positive (NaN/37.9) | Positive (NaN/38.3) | NA | Positive | False negative |
| Symptomatic | Negative | Positive (37.1/36.5) | Positive (NaN/37.0) | NA | Positive | False negative^b^ |
| Symptomatic | Negative | Positive (37.0/36.5) | Positive (NaN/36.6) | NA | Positive | False negative |
| **Nasopharyngeal** | | | | | | |
| Asymptomatic | Positive (35.3) | Negative | Negative | NA | Negative | False positive |
| Asymptomatic | Positive (34.7) | Positive (37.6/NaN) | Negative | Negative (288) | Negative | False positive |
| Asymptomatic | Negative | Positive (NaN/35.9) | Positive (NaN/39.7) | NA | Positive | False negative |
| Symptomatic | Positive (37.1) | Positive (36.7/35.5) | Negative | Negative (286) | Negative | False positive |
| Symptomatic | Positive (36.7) | Negative | Negative | NA | Negative | False positive |
| Symptomatic | Positive (34.3) | Negative | Positive (NaN/36.5) | Negative (274) | Negative | False positive |
| Symptomatic | Negative | Positive (NaN/35.2) | Positive (NaN/36.3) | NA | Positive | False negative^c^ |
| Symptomatic | Negative | Positive (36.1/34.6) | Positive (NaN/36.2) | NA | Positive | False negative^c^ |
| Symptomatic | Negative | Positive (NaN/35.8) | Positive (35.5/38.6) | NA | Positive | False negative |
| Symptomatic | Negative | Positive (36.6/NaN) | Positive (NaN/38.2) | NA | Positive | False negative |
| Symptomatic | Negative | Positive (NaN/34.1) | Positive (NaN/36.5) | NA | Positive | False negative |

Abbreviations: Ct, cycle threshold; CTM, centralized testing method; HCW, healthcare worker; NA, not applicable; NaN, not a number (i.e., negative curve call); RLU, relative light units; SARS-CoV-2, severe acute respiratory syndrome coronavirus 2.

^a^The Cobas^®^ SARS-CoV-2 & Influenza A/B nucleic acid test for use on the Cobas Liat^®^ system is herein referred to as the POC SARS-CoV-2 & Influenza A/B test. The Cobas SARS-CoV-2 & Influenza A/B qualitative assay for use on the Cobas 6800/8800 systems is herein referred to as the 6800/8800 SARS-CoV-2 & Influenza A/B test. Cobas SARS-CoV-2 qualitative assay for use on the Cobas 6800/8800 systems is herein referred to as the 6800/8800 SARS-CoV-2 test.

^b^Of the five false-negative nasal swab samples, three were retested and two were found to be positive after exploratory discordant retesting with the POC SARS-CoV-2 & Influenza A/B test.

^c^Of the six false-negative nasopharyngeal swab samples, two were found to be positive after exploratory discordant retesting with the POC SARS-CoV-2 & Influenza A/B test.

###### Supplemental Table F: Diagnostic performance for the detection of influenza A using the 6800/8800 SARS-CoV-2 & Influenza A/B test compared with the POC SARS-CoV-2 & Influenza A/B test^a^

|  | **6800/8800 SARS-CoV-2 & Influenza A/B test (+) and POC SARS-CoV-2 & Influenza A/B test (+)** | **6800/8800 SARS-CoV-2 & Influenza A/B test (+) and POC SARS-CoV-2 & Influenza A/B test (-)** | **6800/8800 SARS-CoV-2 & Influenza A/B test (-) and POC SARS-CoV-2 & Influenza A/B test (+)** | **6800/8800 SARS-CoV-2 & Influenza A/B test (-) and POC SARS-CoV-2 & Influenza A/B test (-)** | **OPA %**  **(95% CI)** | **PPA %**  **(95% CI)** | **NPA %**  **(95% CI)** |
| --- | --- | --- | --- | --- | --- | --- | --- |
| **Nasal** | | | | | | | |
| Total | 57 | 1 | 1 | 1,149 | 99.8  (99.4, 100.0) | 98.3  (90.9, 99.7) | 99.9  (99.5, 100.0) |
| *Sample type* | | | | | | | |
| Prospective | 22 | 0 | 1 | 999 | 99.9  (99.4, 100.0) | 100.0  (85.1, 100.0) | 99.9  (99.4, 100.0) |
| Retrospective | 35 | 1 | 0 | 150 | 99.5  (97.0, 99.9) | 97.2  (85.8, 99.5) | 100.0  (97.5, 100.0) |
| *Collection method* | | | | | | | |
| HCW-collected | 44 | 1 | 1 | 588 | 99.7  (98.9, 99.9) | 97.8  (88.4, 99.6) | 99.8  (99.0, 100.0) |
| Self-collected | 13 | 0 | 0 | 561 | 100.0  (99.3, 100.0) | 100.0  (77.2, 100.0) | 100.0  (99.3, 100.0) |
| **Nasopharyngeal** | | | | | | | |
| Total | 62 | 2 | 3 | 1,132 | 99.6  (99.0, 99.8) | 96.9  (89.3, 99.1) | 99.7  (99.2, 99.9) |
| Prospective | 19 | 1 | 2 | 1,001 | 99.7  (99.1, 99.9) | 95.0  (76.4, 99.1) | 99.8  (99.3, 99.9) |
| Retrospective | 43 | 1 | 1 | 131 | 98.9  (96.0, 99.7) | 97.7  (88.2, 99.6) | 99.2  (95.8, 99.9) |

Abbreviations: CI, confidence interval; HCW, healthcare worker; NPA, negative percent agreement; OPA, overall percent agreement; PPA, positive percent agreement; SARS-CoV-2, severe acute respiratory syndrome coronavirus 2.

^a^The Cobas^®^ SARS-CoV-2 & Influenza A/B nucleic acid test for use on the Cobas Liat^®^ system is herein referred to as the POC SARS-CoV-2 & Influenza A/B test. The Cobas SARS-CoV-2 & Influenza A/B qualitative assay for use on the Cobas 6800/8800 systems is herein referred to as the 6800/8800 SARS-CoV-2 & Influenza A/B test.

###### Supplemental Table G: Discordant results for the detection of influenza A using the 6800/8800 SARS-CoV-2 & Influenza A/B test compared with the POC SARS-CoV-2 & Influenza A/B test^a^

| **POC RT-PCR**  **POC SARS-CoV-2 & Influenza A/B test result**  **(Ct)** | **CTM 1**  **6800/8800 SARS-CoV-2 & Influenza A/B test**  **(Ct)** | **Discordant type** |
| --- | --- | --- |
| **Nasal (self-collected)** | | |
| Positive (36.0) | Negative | False positive |
| Negative | Positive (38.3) | False negative |
| **Nasopharyngeal** | | |
| Positive (32.8) | Negative | False positive |
| Positive (15.5) | Negative | False positive^b^ |
| Positive (34.7) | Negative | False positive |
| Negative | Positive (39.9) | False negative |
| Negative | Positive (19.2) | False negative^c^ |

Abbreviations: Ct, cycle threshold; CTM, centralized testing method; POC, point-of-care; SARS-CoV-2, severe acute respiratory syndrome coronavirus 2.

^a^The Cobas^®^ SARS-CoV-2 & Influenza A/B nucleic acid test for use on the Cobas Liat^®^ system is herein referred to as the POC SARS-CoV-2 & Influenza A/B test. The Cobas SARS-CoV-2 & Influenza A/B qualitative assay for use on the Cobas 6800/8800 systems is herein referred to as the 6800/8800 SARS-CoV-2 & Influenza A/B test.

^b^This sample was tested for exploratory discordant testing and was found to be negative on retest; growth curve analysis found low PCR volume.

^c^Of the two false-negative nasopharyngeal swab samples, one was found to be positive on retest after exploratory discordant testing with the POC SARS-CoV-2 & Influenza A/B test.

###### Supplemental Table H: Diagnostic performance for the detection of influenza B using the 6800/8800 SARS-CoV-2 & Influenza A/B test compared with the POC SARS-CoV-2 & Influenza A/B test^a^

|  | **6800/8800 SARS-CoV-2 & Influenza A/B test (+) and POC SARS-CoV-2 & Influenza A/B test (+)** | **6800/8800 SARS-CoV-2 & Influenza A/B test (+) and POC SARS-CoV-2 & Influenza A/B test (-)** | **6800/8800 SARS-CoV-2 & Influenza A/B test (-) and POC SARS-CoV-2 & Influenza A/B test (+)** | **6800/8800 SARS-CoV-2 & Influenza A/B test (-) and POC SARS-CoV-2 & Influenza A/B test (-)** | **OPA %**  **(95% CI)** | **PPA %**  **(95% CI)** | **NPA %**  **(95% CI)** |
| --- | --- | --- | --- | --- | --- | --- | --- |
| **Nasal** | | | | | | | |
| Total | 32 | 0 | 0 | 1,176 | 100.0  (99.7, 100.0) | 100.0  (89.3, 100.0) | 100.0  (99.7, 100.0) |
| *Sample type* | | | | | | | |
| Prospective | 0 | 0 | 0 | 1,022 | 100.0  (99.6, 100.0) | NC | 100.0  (99.6, 100.0) |
| Retrospective | 32 | 0 | 0 | 154 | 100.0  (98.0, 100.0) | 100.0  (89.3, 100.0) | 100.0  (97.6, 100.0) |
| *Collection method* | | | | | | | |
| HCW-collected | 32 | 0 | 0 | 602 | 100.0  (99.4, 100.0) | 100.0  (89.3, 100.0) | 100.0  (99.4, 100.0) |
| Self-collected | 0 | 0 | 0 | 574 | 100.0 (99.3, 100.0) | NC | 100.0 (99.3, 100.0) |
| **Nasopharyngeal** | | | | | | | |
| Total | 22 | 0 | 0 | 1,174 | 100.0  (99.7, 100.0) | 100.0  (85.1, 100.0) | 100.0  (99.7, 100.0) |
| Prospective | 0 | 0 | 0 | 1,023 | 100.0  (99.6, 100.0) | NC | 100.0  (99.6, 100.0) |
| Retrospective | 22 | 0 | 0 | 151 | 100.0  (97.8, 100.0) | 100.0  (85.1, 100.0) | 100.0  (97.5, 100.0) |

Abbreviations: CI, confidence interval; HCW, healthcare worker; NC, not calculable; NPA, negative percent agreement; OPA, overall percent agreement; PPA, positive percent agreement; SARS-CoV-2, severe acute respiratory syndrome coronavirus 2.

^a^The Cobas^®^ SARS-CoV-2 & Influenza A/B nucleic acid test for use on the Cobas Liat^®^ system is herein referred to as the POC SARS-CoV-2 & Influenza A/B test. The Cobas SARS-CoV-2 & Influenza A/B qualitative assay for use on the Cobas 6800/8800 systems is herein referred to as the 6800/8800 SARS-CoV-2 & Influenza A/B test.

###### Supplemental Table I: Operator post-study ease of use satisfaction questionnaire results

| **Statement** | **Average agreement with statement score^a^ (1 = Strongly disagree, 5 = Strongly agree)** |
| --- | --- |
| The instructions to add lot and perform controls were easy to follow | 4.1 |
| The instructions to test specimens were easy to follow | 4.5 |
| It was easy to load the sample into the Liat assay tube | 4.6 |
| It was easy to start the assay on the Liat analyzer | 4.7 |
| It was easy to read the test results | 4.8 |
| It was easy to understand the test results | 4.8 |
| The Instructions For Use and Quick Reference Instructions clearly explain what to do if a test result is invalid | 4.2 |
| I did not need help when I tested samples using the Liat assay | 4.4 |
| Overall score | 4.5 |

^a^Statements were scored as follows: 1, strongly disagree; 2, disagree; 3, neutral; 4, agree; 5, strongly agree.
